# Barriers and enablers to diabetic eye screening: a cross sectional survey of young adults with type 1 and type 2 diabetes in the UK

**DOI:** 10.1101/2022.05.24.22275352

**Authors:** Louise Prothero, Martin Cartwright, Fabiana Lorencatto, Jennifer M Burr, John Anderson, Philip Gardner, Justin Presseau, Noah Ivers, Jeremy M Grimshaw, John G Lawrenson, the EROS Study Investigators

**Author notes:** **Corresponding author:** Professor JG Lawrenson, School of Health Sciences, City, University of London, Northampton Square London, EC1V OHB, UK. **Competing interests:** None. **Funder:** This report is independent research funded by the National Institute for Health Research (Policy Research Programme, Enabling diabetic RetinOpathy Screening: Mixed methods study of barriers and enablers to attendance (EROS study), PR-R20-0318-22001). The views expressed in this publication are those of the author(s) and not necessarily those of the NHS, the National Institute for Health Research or the Department of Health and Social Care. **Author contributor statement:** LP, JGL, FL, MC, JMB, PG, JP, NI, JMG conceived the study idea, designed the study and directed the overall conduct of the study; LP/JGL conducted the analysis. LP/JGL wrote the first and final drafts of the paper. All authors provided critical comments, and approved the final version.

## Abstract

**Introduction:** Diabetic retinopathy screening (DRS) attendance in young adults is consistently below recommended levels. The aim of this study was to identify barriers and enablers of diabetic retinopathy screening (DRS) attendance amongst young adults (YA) in the UK living with type 1 (T1D) and type 2 (T2D) diabetes.

**Research design and methods:** YAs (18-34yrs) were invited to complete an anonymous online survey in June 2021 assessing agreement with 30 belief statements informed by the Theoretical Domains Framework of behaviour change (TDF) describing potential barriers/enablers to DRS.

**Results:** In total 102 responses were received. Most had T1D (65.7%) and were regular attenders for DRS (76.5%). The most salient TDF domains for DRS attendance w*ere ‘Goals’*, with 93% agreeing that DRS was a high priority and *‘Knowledge’*, with 98% being aware that screening can detect eye problems early.

Overall 67.4% indicated that they would like greater appointment flexibility [*Environmental context/resources*] and 31.3% reported difficulties getting time off work/study to attend appointments [*Environmental Context/Resources*]. This was more commonly reported by occasional non-attenders versus regular attenders (59.1% vs 23.4%, P=0.002) Most YAs were worried about diabetic retinopathy (74.3%), anxious when receiving screening results (63%) [*Emotion*] and would like more support after getting their results (66%) [*Social influences*]. Responses for T1D and T2D were broadly similar, although those with T2D were more likely have developed strategies to help them to remember their appointments (63.6% vs 37.9%, P=0.019) [*Behavioural regulation*].

**Conclusions:** Attendance for DRS in YAs is influenced by complex interacting behavioural factors. Identifying modifiable determinants of behaviour will provide a basis for designing tailored interventions to improve DRS in YAs and prevent avoidable vision loss.

**Significance of this study:** *What is already known about this subject?:*

- Younger adults (<35 years) with diabetes have been identified as having longer time intervals before attending initial diabetic retinopathy screening (DRS) and are more likely to miss successive screening appointments.
- Previous studies have explored modifiable influences on DRS attendance, but often do not differentiate between population groups, particularly young adults.

*What are the new findings?:*

- One of the main reported barriers to attending DRS was the lack of appointment flexibility and difficulty getting time off work/study to attend appointments. This was compounded by the lack of integration of DRS with other diabetes appointments.
- Most young adults were worried about diabetic retinopathy, anxious when receiving screening results and would like more support

*How might these results change the focus of research or clinical practice?:*

- A more tailored approach is needed to support young adults to attend DRS. The findings of this research provide a basis for developing tailored interventions to increase screening uptake in this age group

## Introduction

Despite evidence supporting the effectiveness of diabetic retinopathy screening (DRS) in reducing the risk of sight loss, attendance for screening in particular demographic groups is consistently below recommended levels.[1] Understanding modifiable barriers and enablers to DRS is essential to develop tailored intervention strategies to improve screening uptake. There have been many studies internationally that have investigated the factors influencing DRS attendance. [2] [3] Barriers/enablers to attendance potentially operate at different levels, including the person with diabetes, the healthcare professional or the healthcare system. Furthermore, factors influencing individual screening attendance are likely to differ according to the presence of variables that are known to impact on health equity, e.g. type of diabetes, ethnicity or socioeconomic status. [4-7] However, studies have often considered people with diabetes as a homogenous group and relatively few studies have addressed barriers/enablers in particular population subgroups.

One demographic group where adherence to DRS consistently falls below recommended levels is young adults (YAs) with diabetes aged under 35 years. [8-11] Recent studies from the UK Diabetic Eye Screening Programme (DESP) have shown that the time interval from registration with the screening programme to DRS attendance is significantly longer for the 18–34-year age group, with approximately 20% remaining unscreened three years after registration. [10] Furthermore, younger adults (<35 years) are more likely to miss three successive DRS appointments. [11]

This is a particularly hard to reach group and there has been little previous research to understand the reasons for poor DRS attendance in YAs. [12] A 2017 Australian study [13] conducted semi structured interviews with YAs, N=10 aged 18–39 years and older adults, N=20 aged over 40 years with type 2 diabetes (T2D). This study utilised a behavioural science framework, the Theoretical Domains Framework [TDF]), [14] to explore the wide range of barriers and enablers to attendance. The TDF synthesises constructs from 33 theories of behaviour change into 14 domains, representing individual, socio-cultural and environmental influences on behaviour (e.g. knowledge, emotions, social and professional identity, perceived consequences, intention, environmental context and resources). Although younger and older adults shared several screening behaviour determinants, a number of TDF domains showed greater salience to YAs including: misconceptions regarding diabetic retinopathy [*Knowledge*]; social comparison with others [6]; unrealistic optimism and perceived invulnerability [*Beliefs about consequences*]; and lack of time and financial resources [*Environmental context and resources*] [13]. We have recently completed the NIHR-funded ‘Enabling diabetic RetinOpathy Screening: Mixed methods study of barriers and enablers to attendance (EROS study)’, which aimed to identify barriers and enablers to DRS attendance experienced by YAs with diabetes living in the UK. A part of this research we conducted qualitative interviews with 29 YAs with type 1 diabetes (T1D) aged 18-34 years.[15] We similarly applied the TDF to identify modifiable barriers and enablers to DRS attendance. Key influences fell within the TDF domains: [*Knowledge*] e.g. not understanding reasons for attending DRS or treatments available if diabetic retinopathy is detected; [*Social support*] e.g. lack of support following DRS results; [S*ocial role and Identity*] e.g. not knowing other people their age with diabetes; feeling ‘isolated’ and being reluctant to disclose their diabetes; [*Environmental Context and Resources*] e.g. lack of appointment flexibility and options for rescheduling) and [Emotion] e.g. diabetes distress/burnout. Enablers included: [*Social Influences*] e.g. support of family/diabetes team; and [*Goals*] e.g. DRS regarded as ‘high priority’. Barriers/enablers were generally consistent across groups defined by patterns of attendance (regular attenders, occasional non-attenders, regular non-attenders).

In the current study, we used the results of the previous interview study [15] to design an online survey to assess the generalisability of the perceived barriers and enablers in a more diverse sample of YA with regard to particular demographic characteristics (e.g. age, employment, gender, ethnicity, educational level). We also investigated differences in perceived barriers and enablers between YAs with T1D and T2D and in those that attend DRS regularly versus those who did not. The survey also served to triangulate findings from qualitative and quantitative methods to gain a more complete picture of the factors that influence screening uptake in YAs. [16]

## Research design and methods

### Design

A cross-sectional web-based survey.

### Ethical approvals

This study received ethical approval from the NHS Wales Research Ethics Committee 2 (REC reference: 19/WA/0228). Prior informed consent was obtained from all participants.

### Participants and recruitment strategy

Eligible participants included YAs aged 18-34 years with diabetes. Previous studies have shown that people in this age group are least likely to attend DRS and have high rates of referable retinopathy. [8] [10,11] As this was a descriptive survey, we did not have a pre-defined target sample size in mind and aimed to maximise response rate from as many YAs as possible. Two recruitment strategies were used:

1. A text (SMS) message with the link to the survey was sent to all YAs with T1D and T2D aged 18-34 on the register of a large Diabetic Eye Screening Programme (DESP) in London whose mobile number was available. Screening providers often use this mobile phone strategy to request feedback from patients about the care they receive.
2. The survey was also promoted via the web pages of the Juvenile Diabetes Research Foundation UK (JDRF UK) and Diabetes UK and further supported using the Facebook and Twitter accounts of these organisations (see Supplementary file S1 for examples of promotional material).

### Materials: Questionnaire

The full survey is available in Supplementary file S2. In brief, the survey was developed based on guidance for conducting surveys using the TDF[17] and the findings of our previous interview study with YAs in the UK. [15] The survey was fully anonymous and divided into three sections:

- Section 1 Participant demographics: age, gender, ethnicity, geographical location, highest level of education; type and duration of diabetes; screening appointments missed in the last 3 years (either forgotten and rescheduled or deliberately not attended) (12 questions).
- Section 2 Perceived influences on DRS attendance: Participants were presented with 30 belief statements representing barriers and enablers to DRS attendance. These statements were developed based on the inductively generated themes based on frequency and elaboration from our semi-structured interview study with young adults in the UK [15](e.g., the theme ‘*Diabetic retinopathy is a concern’* was reflected in the belief statement ‘I *worry about diabetic retinopathy’*)). To ensure theoretical coverage and that the wide range of potential influences were considered, belief statements covered 13 of the 14 TDF domains (our earlier qualitative study [15] did not identify themes for the domain *Optimism*). Participants rated their agreement with each statement using a 5-point Likert scale (strongly agree → strongly disagree).
- Section 3: Free text question ‘Please describe any other factors which influence your attendance at diabetic eye screening which we have not covered’

To assess participant burden, clarity of questions and face validity, a draft questionnaire was sent to the project Patient and Public Involvement (PPI) panel consisting of four YAs with diabetes, who were asked to comment on the following:

- How long the survey takes to complete
- If the survey items make sense, are appropriate and relevant
- If there are any survey items which are ambiguous or unclear

A final version based on the feedback was uploaded and pre-tested for technical quality prior to distribution.

### Procedure

The survey took place in June 2021. The questionnaire was hosted online using Qualtrics Survey Software (https://www.qualtrics.com/uk/core-xm/survey-software/). The survey was fully anonymous, and participants consented to participate in the survey by completing a brief consent form on the survey home page. Respondents were offered an incentive in the form of the chance to win a £20 Love2shop voucher (we offered twenty £20 vouchers).

### Analysis

After the closure of the survey, all data were imported into an Excel spreadsheet. Data were summarised using descriptive statistics (percentages [n)). For the responses to the 5-point scale in Section 2, scores for ‘Strongly agree’ and ‘Somewhat agree’ were combined into an overall mean and percentage agreement score.

Statistical analysis for comparison between participants with T1D and T2D and pattern of attendance (regular attenders versus occasional non-attenders), was carried out using MedCalc Statistical Software version 18 (MedCalc Software, Ostend, Belgium; http://www.medcalc.org) in the form of Chi-squared tests to determine differences in endorsed barriers and enablers. Occasional non-attenders were grouped as those who had either (a.) unintentionally forgotten/missed previous appointments and rescheduled, or (b.) actively chosen to not attend on at least one occasion.

Free text survey responses were coded thematically by a single investigator (JGL) to identify common themes and concepts.

## Results

### Respondent Demographic Characteristics

One hundred and two responses were received. Detailed respondent demographic characteristics are presented separately for respondents with T1D and T2D in Table 1 (two participants reported having an ‘other’ type of diabetes (e.g. maturity onset diabetes of the young (MODY)). Almost all respondents (98.5%) were located in England. The greatest proportion of respondents were female (59.8%), had T1D (65.7%), were aged 30-34 (52.9%), identified as White British (57.8%), employed (76.4%) and educated to degree level or higher (60.8%).

**Table 1:** Characteristics of persons with type 1 and type 2 diabetes

|  | **Type 1**<br>**N=67** | **Type 2**<br>**N=33** |
| --- | --- | --- |
| **Gender** | N (%) | N (%) |
| Male | 20 (29.9) | 17 (51.5) |
| Female | 45 (67.2) | 16 (48.5) |
| Other | 2 (3.0) | 0 (0.0) |
| **Age** |  |  |
| 18 -23 | 13 (19.4) | 1 (3.0) |
| 24-29 | 25 (38.8) | 5 (15.2) |
| 30-34 | 28 (41.8) | 25 (75.8) |
| NR | 0 (0.0) | 2 (6.1) |
| **Number of years since diagnosis** |  |  |
| <3 | 4 (1.5) | 11 (33.3) |
| 3 to 9 | 18 (28.4) | 19 (57.6) |
| 10 to 14 | 12 (17.9) | 3 (9.1) |
| 15 to 19 | 15 (20.9) | 0 (0.0) |
| >20 | 16 (23.9) | 0 (0.0) |
| NR | 1 (1.5) | 0 (0.0) |
| **Ethnicity** |  |  |
| White British | 48 (71.6) | 10 (30.3) |
| Other white background | 8 (11.9) | 0 (0.0) |
| Indian | 3 (4.5) | 8 (24.2) |
| Bangadeshi | 1 (1.5) | 6 (18.2) |
| Pakistani | 1 (1.5) | 4 (12.1) |
| Other | 4 (6.0) | 4 (12.1) |
| NR | 2 (3.0) | 1 (3.0) |
| **Country of residence** |  |  |
| England | 66 (98.5) | 33 (100.0) |
| Wales | 1 (1.5) | 0 (0.0) |
| **Area** |  |  |
| Urban | 31 (46.3) | 18 (54.5) |
| Suburban | 35 (52.2) | 14 (42.4) |
| Rural | 1 (1.5) | 0 (0.0) |
| NR | 0 (0.0) | 1 (3.0) |
| **Occupational Status** |  |  |
| Full time job | 46 (68.7) | 18 (54.5) |
| Part time job | 7 (10.4) | 3 (9.1) |
| Self-employed | 2 (3.0) | 2 (6.1) |
| Studying | 6 (9.0) | 2 (6.1) |
| Unemployed | 4 (6.0) | 5 (15.2) |
| Other/NR | 2 (3.0) | 3 (9.1) |
| **Highest level of education** |  |  |
| Secondary | 3 (4.5) | 7 (21.2) |
| Further Education | 16 (23.9) | 6 (18.2) |
| Degree or higher | 45 (67.2) | 17 (51.5) |
| Other/NR | 3 (4.5) | 3 (9.1) |
| **Missed DRS appointments** |  |  |
| None | 51 (76.1) | 27 (81.8) |
| Unintentionally | 14 (20.9) | 6 (18.2) |
| Intentionally | 2 (3.0) | 0 (0.) |

Most respondents were regular attenders for eye screening, with 76.5% having not missed a DRSs appointment in the last three years.

### Barriers and enablers

The results of agreement/disagreement with belief statements related to barriers/enablers to diabetic retinopathy screening are presented in Table 2, and Figure 1. In this well-engaged population, the most salient TDF domain for DRS attendance was *Goals*, with 93% of respondents agreeing that DRS was a high priority in terms of diabetes management, with a clear intention to attend future eye screening appointments [TDF domain, *Intention*] (97%). The reason for attending was understood by almost all respondents, with 98% being aware that diabetic eye screening can help detect eye problems early [*‘Knowledge’*]. Conversely, 84% did not know what treatments were available if retinopathy was detected [*Knowledge*]. Importantly, only 52% felt that diabetes education and training covered eye screening in detail [*Skills*].

**Table 2.**
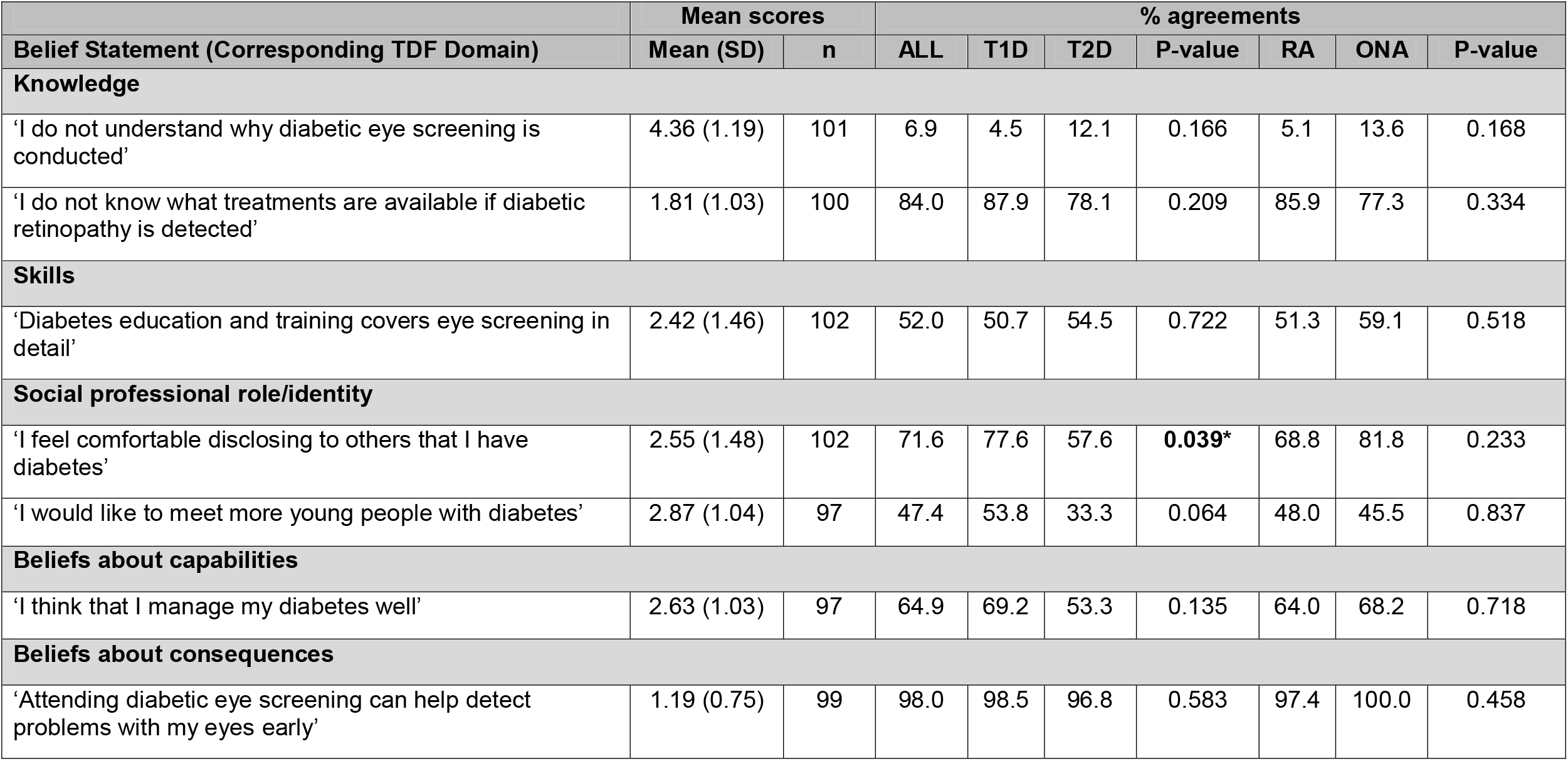
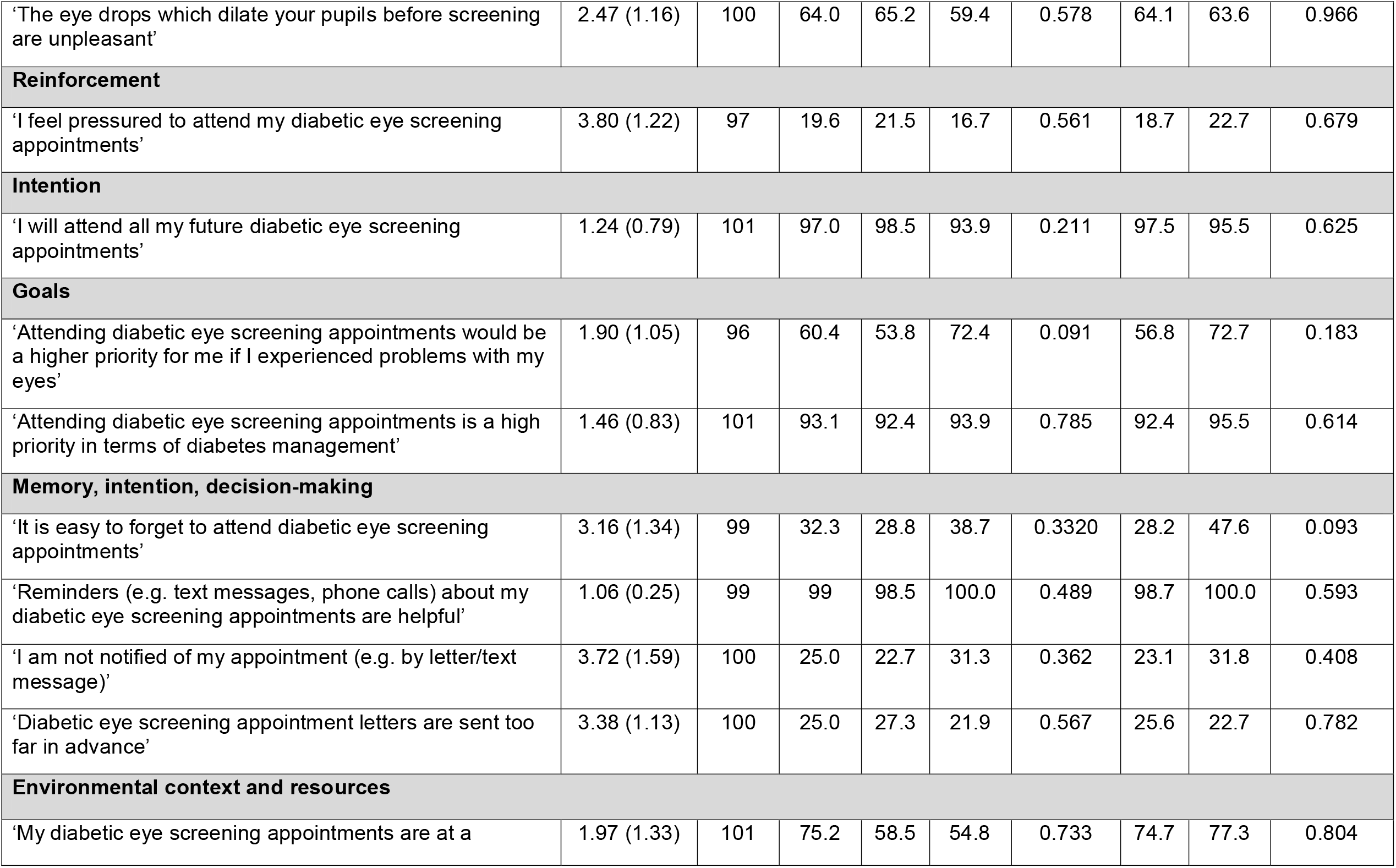
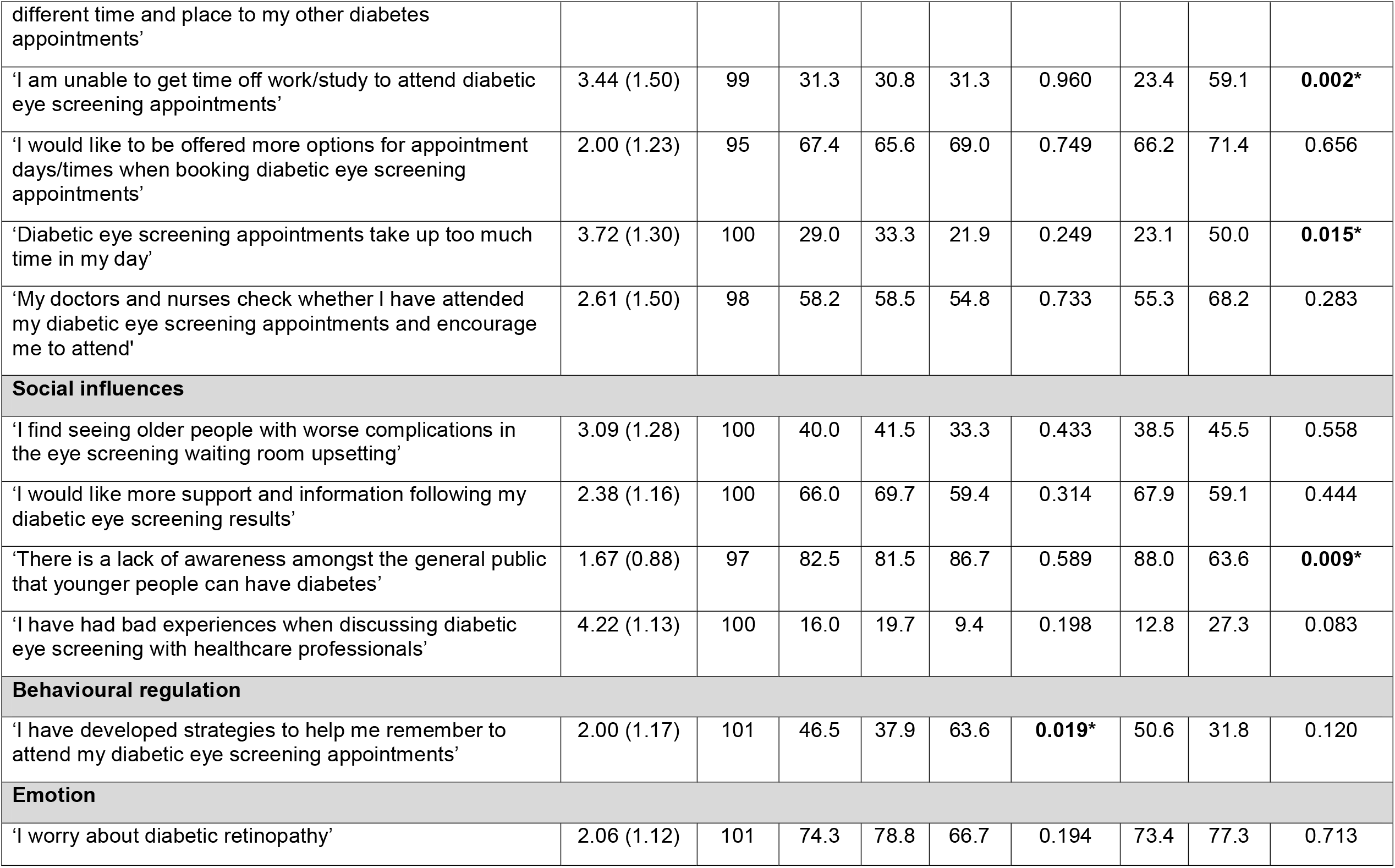
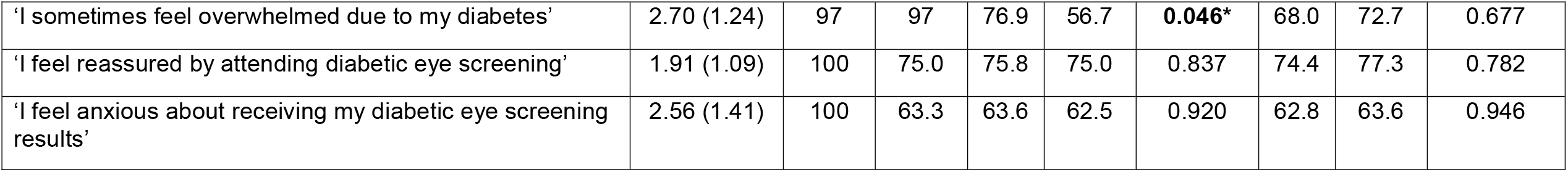
Mean scores and percentage agreement with belief statements representing barriers and enablers to diabetic retinopathy screening. The mean scores correspond to the extent to which participants agreed with each statement using a 5-point Likert scale (strongly agree=1; somewhat agree=2; neither agree nor disagree=3; somewhat disagree=4; strongly disagree=5). The p-value represents the results of the Chi-square test for differences between type 1 (T1D) type 2 diabetes (T2D) sub-groups and between regular attenders (RA) and occasional non-attenders (ONA). Highlighted values indicate statistically significant differences

**Figure 1.**
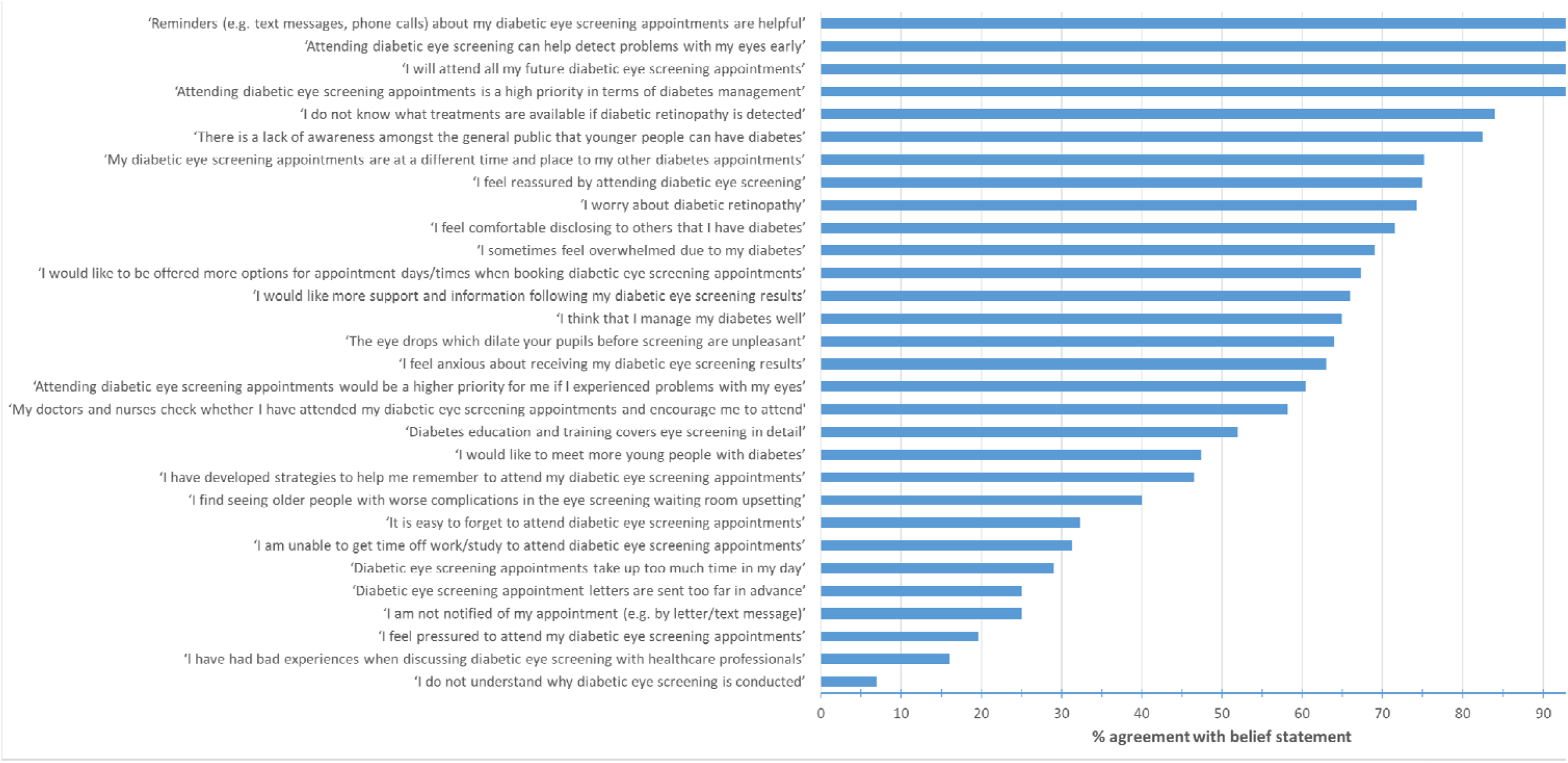
Rank order of percentage agreement with each belief statements for all survey respondents

In terms of the screening process itself, overall 67.4% indicated that they would like to be offered more options for appointment days/times when booking diabetic eye screening appointments [*Environmental context/resources*] and approximately one third of respondents had difficulties getting time off work/study to attend diabetic eye screening appointments [*Environmental Context/Resources]*. Scheduling was compounded by retinopathy screening appointments being at a different time to other diabetes appointments for most respondents [*Environmental context/resources*] (75.2%). Comparison between regular attenders and occasional non-attenders (Table 2) found that non-attenders were more likely to report more difficulty taking time off to attend appointments (59.1% vs 23.4%, P=0.002) and felt that appointments took up too much of the day 50.0% vs 23.1%, P=0.015).

Additional negative aspects of the screening process were acknowledged e.g. being upset by seeing older people with worse complications in the waiting room [*Emotion*] (40%) and the adverse effects of the dilating eye drops [*Beliefs about consequences*](64%). Most respondents were worried about diabetic retinopathy (74.3%), reported anxiety when receiving screening results [*Emotion]* (63%) and would like more support and information after getting their results [*Social influences* (66%).

Enablers of attendance included an awareness that DRS allows early detection of eye problems [*Beliefs about consequences*] (98%), feeling reassured by attending DRS [*Emotion*} (75%) and feeling comfortable disclosing diabetes to others [*Social identity*] (71.6%).

Percentage agreement between YAs with T1D and T2D were broadly similar (Table 2). Statistically significant differences were found for only three statements. Persons with T1D were more likely to feel overwhelmed by their diabetes (76.9% vs 56.7%, P=0.046) [*Emotion*]. YAs with T2D were less comfortable in disclosing their diabetes to others (77.6% vs 57.6%, P=0.039) [*Social identity*] and were more likely have developed strategies to help them to remember to attend their DRS appointments (63.6% vs 37.9%, P=0.019) [*Behavioural regulation*].

### Other factors influencing attendance at diabetic eye screening

Free text responses were received from 30 respondents. These covered the following areas: impact of the Covid pandemic on scheduling DRS appointments (‘*Last appointment later due to Covid’*); fear of vision loss (*‘My mother In law lost her eye due to diabetic (retinopathy)’*); appointment inflexibility (‘*They can also be inflexible-the nearest one to me only does Tuesday mornings’*); impact of eye drops and transport issues (‘*They can be difficult to get to, especially as you cannot drive-in one occasion I had to get 3 buses which took nearly 2 hours-the return trip with dilated pupils wasn’t fun’)*, interactions with screening staff and issues with receiving screening results (*‘I always receive letters that are extremely distressing and usually on a weekend when I cannot call anyone’*). The complete set of free text comments can be found in the supplementary material.

## Discussion

### Summary of findings

This study aimed to build on the findings of our earlier qualitative interview study to identify perceived barriers and enablers to DRS in YAs in the UK. [15] The results broadly confirmed our qualitative interview findings in YAs with T1D and converged with the findings from a qualitative study of YAs with T2D in Australia. [13]

Based on the level of agreement with each belief statement, the most salient TDF domains associated with DRS included ‘Social influences’, ‘Intentions’, ‘Emotion’, ‘Environmental context/resources’, ‘Knowledge’, ‘Skills’, and ‘Goals’. However, the level of agreement with belief statements differed between respondents with T1D and T2D and between regular attenders and occasional non-attenders. This emphasises the importance of understanding barriers/enablers for specific population sub-groups.

Overall, survey respondents represented a well-engaged population, with approximately 78% reporting that they had not missed any screening appointments in the last 3 years. This may explain the high level of understanding of the purpose of DRS, the high priority (*Goals*) given to this particular aspect of diabetes care and the strong intention to attend further screening appointments. However, despite a large majority understanding the purpose of DRS and the need for regular screening, there were specific knowledge gaps such as an awareness of the treatments available should sight-threatening retinopathy be detected.

Competing time demands and practical issues with making appointments have been previously shown to be important barriers to DRS attendance. [2,7,13,15] This was particularly pertinent amongst respondents who reported occasionally missing DRS appointments. Occasional non-attenders were more likely to agree with the belief statements: ‘*Diabetic eye screening appointments take up too much time in my day*’ and ‘*I am unable to get time off work/study to attend diabetic eye screening appointments*’. Most survey respondents agreed that their DRS appointments were at a different time and place to their other diabetes appointments. Possible solutions could include evening and weekend appointments coupled with a flexible online booking system to allow them to schedule appointments more easily. Furthermore, the use of an integrated diabetes care in the form of a ‘one-stop shop’ clinic that combines several processes of diabetes care, including DRS, which would reduce the number of appointments. Although there is currently no high quality evidence from the UK that integrated diabetes clinics improves DRS uptake specifically in YA, ‘collaborative case management’, which coordinates processes of diabetes care, have been shown to improve diabetic retinopathy outcomes in clinical trials of a general adult population with diabetes. [1]

The eye drops used to temporarily dilate the pupils were perceived to be unpleasant by the majority respondents. Free text comments also alluded to the impact of the eye drops on attendance due to not being able to drive to and from the screening venue and having to rely on family members or use public transport (‘*With the drops you can’t drive but it’s also hard to then see where the train is. The drops knock me out for the rest of the day and really affect work and everything’*). Previous studies have also reported on barriers relating to dilating eye drops [18]. The National Screening committee (NSC) in the UK currently recommends pupil dilation (mydriasis) for all attendees for DRS based on the ease of organisation and improvement in image quality, however there is evidence that using mydriasis only when clinically necessary can be effective for DRS. [19]

Fear of diabetic retinopathy was identified as a cause for concern, with a high level of agreement that screening attendance provided reassurance. However, there was a particular anxiety associated with receiving screening results and a desire for more support and information on receiving results. The previous literature identified that whilst, for some, the fear of losing vision is a strong incentive to attend DRS, for others, the fear of a diagnosis of diabetic retinopathy may act as a barrier [18]. Interventions to address this could include training suitably qualified screeners to give immediate feedback on the results of eye screening or the provision of more support after receiving results. This strategy is potentially acceptable to implement in practice, with a recent cross sectional survey of healthcare professionals working in the UK National Diabetic Eye Screening Programme highlighting that screening providers would like to be more involved in discussing screening results with YAs and promoting diabetes self-management.[20]

A recommendation by a healthcare professional (HCP) has been shown to be an important enabler for DRS uptake and receiving a recommendation from a healthcare provider to attend screening is associated with improved attendance. [2,13] However, survey respondents reported that members of the diabetes team did not always check their DRS attendance record or encourage them to attend. This is clearly a missed opportunity to improve screening uptake in this population.

T2D is increasingly prevalent in YAs. This trend is particularly pronounced in South Asian ethnic groups.[21,22] Previous research has established that YAs with T2D are at higher risk of developing diabetic retinopathy, [23,24] face unique barriers to diabetes self-management and have specific unmet psychosocial needs. [25] Although all participants in our earlier interview study [15] had T1D, 32% of respondents in the current study were YAs with T2D. The levels of agreement between these sub-groups were very similar, with significant differences in agreement found for only three belief statements (*‘I sometimes feel overwhelmed due to my diabetes’*; ‘*I feel comfortable disclosing to others that I have diabetes’; ‘I have developed strategies to help me remember to attend my diabetic eye screening appointments’*). Respondents with T1D were more likely to feel overwhelmed by their diabetes. YAs with type 2 diabetes were less comfortable in disclosing their diabetes to others. Research in Australia suggests that young people with T2D are sensitive to stigmatising attitudes. [26] Although there was no difference between T1D and T2D respondents in terms of likelihood of forgetting to attend DRS appointments, T2D respondents were more likely to have developed strategies to help them to remember to attend.

### Strengths and limitations of the current study

One of the strengths of the current study is that it addresses an important evidence gap. Although there are many studies that have reported modifiable barrier/enablers to DRS [2,3], the majority of these studies tend to treat people with diabetes as a homogeneous group, and therefore, it is not possible to identify determinants of DRS uptake from the perspective of particular population subgroups. Relatively few studies have reported barriers from the perspective of YAs, who are at a high risk of developing sight-threatening retinopathy. [10,23,24]

Another strength of our approach is the use of a theory-informed methodology to identify barriers and enablers. [14] We used the TDF to guide data collection, which provides a basis for generating future behaviour change strategies that can be tailored to YA to address barriers or enhance facilitators.

Although this hypothesis-generating study was limited in terms of its small sample size, we received responses from a demographically diverse sample of YA, including 33% of responses from YA with T2D. The results confirmed many of barriers and enablers identified in previous qualitative interview studies [13,15] and suggest that that the determinants of screening attendance are broadly similar for YAs with T1D and T2D.

The main limitation was the difficulty experienced in recruiting non-attenders. Despite using a variety of recruitment strategies (see Appendix), only 20% of survey respondents had missed a DRS appointment in the last 3 years and nearly all of these had unintentionally missed the appointment (i.e. they forget or were unable to attend). We were only able to recruit two participants who had made a deliberate decision not to attend DRS), which could impact on the generalisability of the findings to repeat non-attenders.

### Conclusions and implications for policy

Barriers identified in the current study included the lack of appointment flexibility, impact of the eye drops used to dilate the pupils and anxiety associated with the risk of developing diabetic retinopathy. More consistent checking of DRS attendance by the diabetes team and encouragement to attend could be an important enabler. These findings highlight recommendations for changing policy and practice, including pinpointing to specific intervention strategies that could potentially address identified barriers and enablers and increase attendance to DRS in this priority population group. Future research should address the challenges of engaging with socially disadvantaged and hard to reach groups to ensure that they are not excluded.

## Data Availability

All data produced in the present study are available upon reasonable request to the authors

## Acknowledgements

The authors wish to acknowledge the help of the following: Diabetes UK; JDRF (the type 1 diabetes research charity); Louis Boulter (DESP lead North East London); EROS PPI panel; EROS Research Advisory Group. JMG holds a Canada Research Chair in Health Knowledge Transfer and Uptake. NMI holds a Canada Research Chair in Implementation of Evidence-Based Practice.

## Supplementary Material

### S1: Rapid Digital Survey. Promotional material

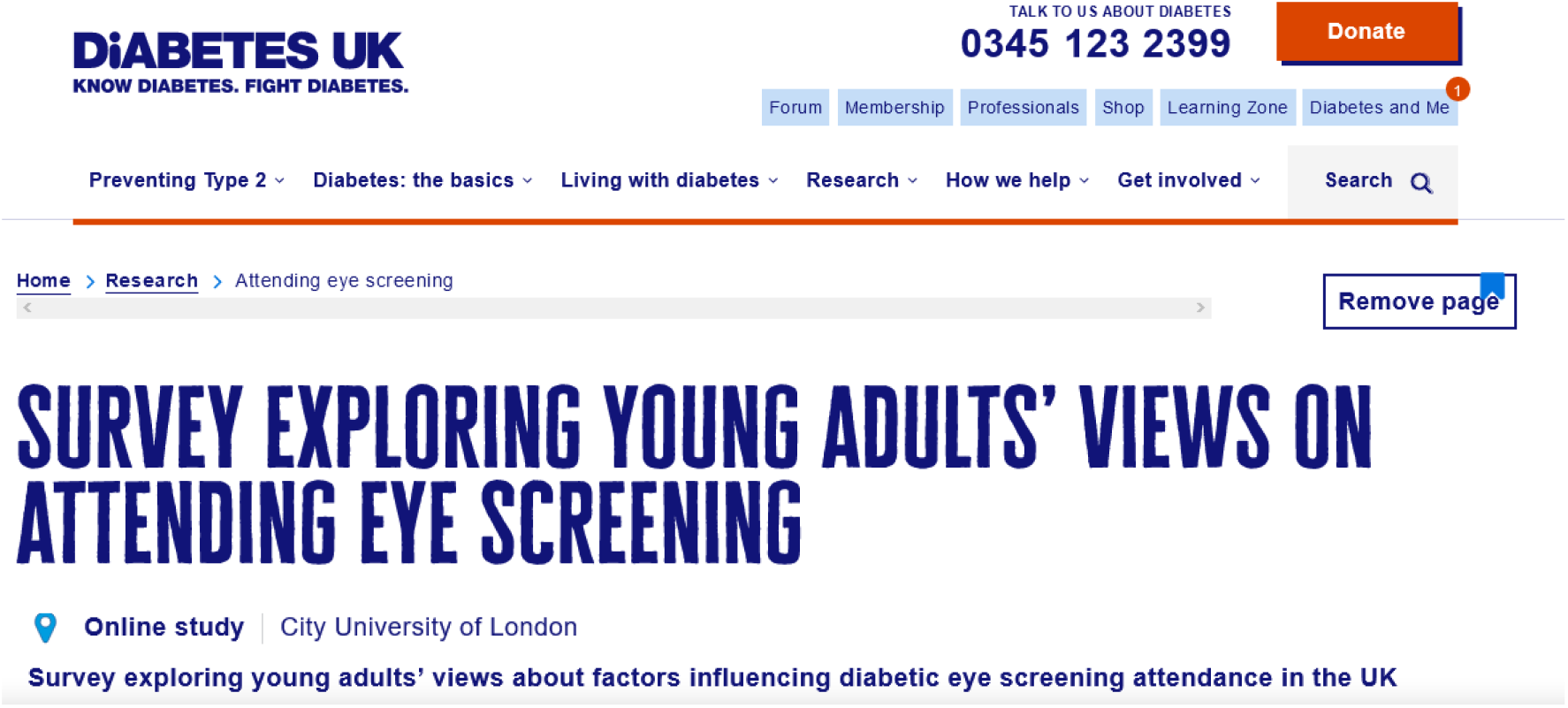

Survey exploring young adults’ views about factors influencing diabetic eye screening attendance in the UK

Researchers at City University of London would like to invite people aged 18-34 years living with diabetes, to complete an online survey that explores a number of issues that may affect their attendance at diabetic eye screening and views on the screening process. The information you provide will help the researchers to make specific recommendations to improve the eye screening service.

The survey will take approximately 10 minutes to complete and you can <u>take part here</u>.

There will be an opportunity to enter a prize draw to win one of twenty Love2shop £20 vouchers.

For more information please contact Professor John Lawrenson at

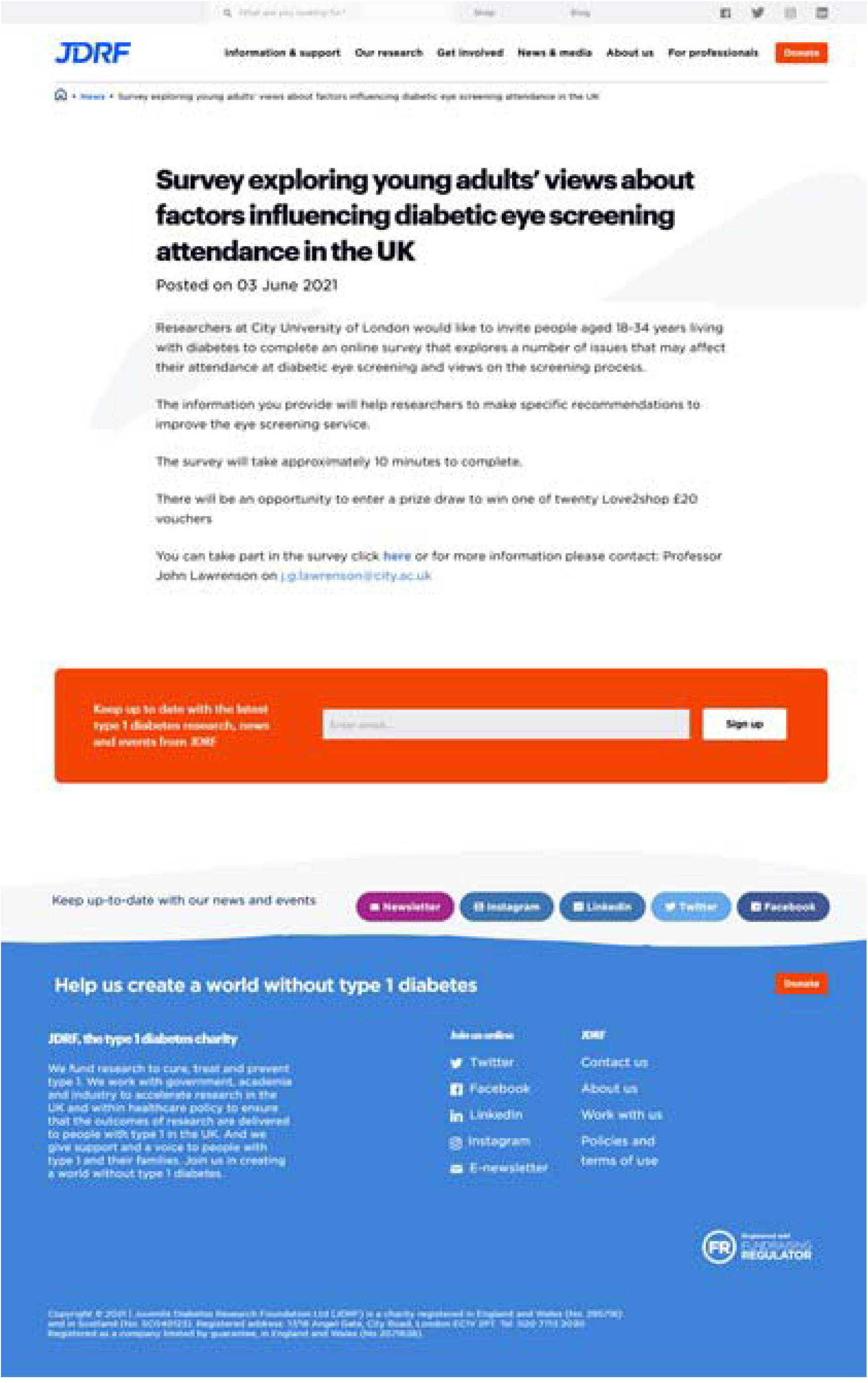

### S2: Copy of Rapid Digital Survey

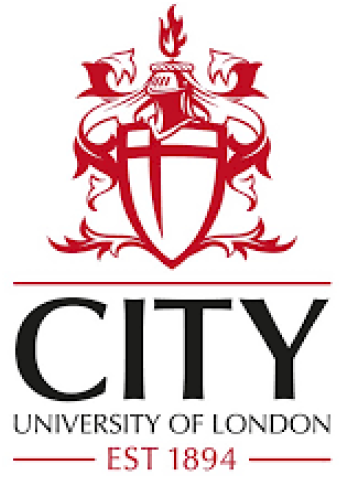

#### Rapid Digital Survey

**The purpose of the survey is to explore young adults’ views about factors influencing diabetic eye screening attendance**. The information you provide will help us to make recommendations in order to improve the eye screening service.

**The survey will take approximately 10 minutes to complete**. Before completing the survey, you will be asked to provide your consent to participate and some information about yourself. The information you provide will be used to describe who took part in our survey as a whole. Everyone’s responses will be combined so you will not be identifiable.

Should you wish to enter the prize draw, you will be asked to provide a contact e-mail address. **Your e-mail address will be separated and stored separately from your survey responses, and will only be used to contact you if you are picked from the prize draw to win a £20 Love2shop voucher. Following the prize draw, all e-mail addresses provided will be deleted**.

This survey is part of a wider study (the EROS study), which has been approved by Wales Research Ethics Committee 2 (REC reference: 19/WA/0228). If you have any questions or concerns regarding completing the survey, please e-mail the principal investigator Prof John Lawrenson

Thank you for taking the time to take part in our survey,

The EROS Study team

○ I understand that my participation is voluntary and that I am free to withdraw at any time without giving a reason and without my legal rights being affected
○ I understand that all data and results from this study will be treated strictly confidential and anonymised, so that no individual can be identified from the data.
○ I understand that data collected about me during this study will be stored on a secure database at City, University of London for use by the research team to enable them to analyse the data
○ I agree to take part in this study

1. **Where do you live:**
  ⍰ England
  ⍰ Northern Ireland
  ⍰ Scotland
  ⍰ Wales
2. **Which of the following best describes the area you live in:**
  ⍰ Urban (e.g. city centre)
  ⍰ Suburban (e.g. residential area on the edge of a large town/city)
  ⍰ Rural (e.g. countryside)
3. **Your age group:**
  ⍰ 18-23 years
  ⍰ 24-29 years
  ⍰ 30-34 years
4. **Gender:**
  ⍰ Male
  ⍰ Female
  ⍰ Other _ _ _ _ _ _ _
  ⍰ Prefer not to say
5. **Ethnicity:**

6. **Highest level of education:**
  ⍰ Secondary school (e.g. GCSEs, National 5s)
  ⍰ Further education (e.g. AS/A-levels, Scottish Higher/Advanced Higher)
  ⍰ Bachelor’s degree or more (e.g. BSc/BA, MSc, PhD)
  ⍰ Other _ _ _ _ _ _ _
  ⍰ Prefer not to say
7. **Main occupation:**
  ⍰ Full-time job
  ⍰ Part-time job
  ⍰ Studying full-time
  ⍰ Studying part-time
  ⍰ Self-employed
  ⍰ Unemployed
  ⍰ Other _ _ _ _ _ _ _
  ⍰ Prefer not to say
8. **Type of diabetes:**
  ⍰ Type 1 diabetes
  ⍰ Type 2 diabetes
  ⍰ If other please specify _ _ _ _ _ _ _
9. **Approximate year you received your diabetes diagnosis:** _ _ _ _ _ _ _
10. **The number of diabetic eye screening appointments I have attended in the last 3 years is** _ _ _ _ _ _ _
11. **The number of diabetic eye screening appointments I have chosen not to attend in the last 3 years is** _ _ _ _ _ _ _
12. **The number of diabetic eye screening appointments I have forgotten and rescheduled in the last 3 years is** _ _ _ _ _ _ _

**5. Ethnicity:**
|  |  |  |  |
| --- | --- | --- | --- |
| A: British | ☐ | J: Bangladeshi | ☐ |
| B: Irish | ☐ | K: Any other Asian background | ☐ |
| C: Any other White background | ☐ | L: Caribbean | ☐ |
| D: White and Black Caribbean | ☐ | M: African | ☐ |
| E: White and Black African | ☐ | N: Any other Black background | ☐ |
| F: White and Asian | ☐ | O: Chinese | ☐ |
| G: Any other mixed background | ☐ | P: Any other ethnic group | ☐ |
| H: Indian | ☐ | Prefer not to say | ☐ |
| I: Pakistani | ☐ |  |  |

Please read each statement below and indicate your agreement by choosing one of five responses:

Strongly agree

Somewhat agree

Neither agree nor disagree

Somewhat disagree

Strongly disagree

[Presented in 3 x blocks of 10]

|  |  |  |  |  |  |
| --- | --- | --- | --- | --- | --- |
| 1. | 'Diabetes education and training covers eye screening in detail' |  |  |  |  |
|  | 1 | 2 | 3 | 4 | 5 |
| 2. | 'I feel comfortable disclosing to others that I have diabetes' |  |  |  |  |
|  | 1 | 2 | 3 | 4 | 5 |
| 3. | 'Attending diabetic eye screening can help detect problems with my eyes early' |  |  |  |  |
|  | 1 | 2 | 3 | 4 | 5 |
| 4. | 'I will attend all my future diabetic eye screening appointments' |  |  |  |  |
|  | 1 | 2 | 3 | 4 | 5 |
| 5. | 'It is easy to forget to attend diabetic eye screening appointments' |  |  |  |  |
|  | 1 | 2 | 3 | 4 | 5 |
| 6. | 'My diabetic eye screening appointments are at a different time and place to my other diabetes appointments' |  |  |  |  |
|  | 1 | 2 | 3 | 4 | 5 |
| 7. | 'I worry about diabetic retinopathy' |  |  |  |  |
|  | 1 | 2 | 3 | 4 | 5 |
| 8. | 'I have developed strategies to help me remember to attend my diabetic eye screening appointments' |  |  |  |  |
|  | 1 | 2 | 3 | 4 | 5 |
| 9. | 'I do not understand why diabetic eye screening is conducted' |  |  |  |  |
|  | 1 | 2 | 3 | 4 | 5 |
| 10. | 'Attending diabetic eye screening appointments is a high priority in terms of diabetes management' |  |  |  |  |
|  | 1 | 2 | 3 | 4 | 5 |
| 11. | 'Reminders (e.g. text messages, phone calls) about my diabetic eye screening appointments are helpful' |  |  |  |  |
|  | 1 | 2 | 3 | 4 | 5 |
| 12. | 'I am unable to get time off work/study to attend diabetic eye screening appointments' |  |  |  |  |
|  | 1 | 2 | 3 | 4 | 5 |
| 13. | 'My doctors and nurses check whether I have attended my eye screening and encourage me to attend ' |  |  |  |  |
|  | 1 | 2 | 3 | 4 | 5 |
| 14. | 'I sometimes feel overwhelmed due to my diabetes' |  |  |  |  |
|  | 1 | 2 | 3 | 4 | 5 |
| 15. | 'There is a lack of awareness amongst the general public that younger people can have diabetes' |  |  |  |  |
|  | 1 | 2 | 3 | 4 | 5 |
| 16. | 'I would like to meet more young people with diabetes' |  |  |  |  |
|  | 1 | 2 | 3 | 4 | 5 |
| 17. | 'I think that I manage my diabetes well' |  |  |  |  |
|  | 1 | 2 | 3 | 4 | 5 |
| 18. | 'I feel pressured to attend my diabetic eye screening appointments' |  |  |  |  |
|  | 1 | 2 | 3 | 4 | 5 |
| 19. | 'Attending diabetic eye screening appointments would be a higher priority for me if I experienced problems with my eyes' |  |  |  |  |
|  | 1 | 2 | 3 | 4 | 5 |
| 20. | 'I would like to be offered more options for appointment days/times when booking diabetic eye screening appointments' |  |  |  |  |
|  | 1 | 2 | 3 | 4 | 5 |
| 21. | 'I find seeing older people with worse complications in the eye screening waiting room upsetting' |  |  |  |  |
|  | 1 | 2 | 3 | 4 | 5 |
| 22. | 'I feel reassured by attending diabetic eye screening' |  |  |  |  |
|  | 1 | 2 | 3 | 4 | 5 |
| 23. | 'I do not know what treatments are available if diabetic retinopathy is detected' |  |  |  |  |
|  | 1 | 2 | 3 | 4 | 5 |
| 24. | 'The eye drops which dilate your pupils before screening are unpleasant' |  |  |  |  |
|  | 1 | 2 | 3 | 4 | 5 |
| 25. | 'I am not notified of my appointment (e.g. by letter/text message)' |  |  |  |  |
|  | 1 | 2 | 3 | 4 | 5 |
| 26. | 'I have had bad experiences when discussing diabetic eye screening with healthcare professionals' |  |  |  |  |
|  | 1 | 2 | 3 | 4 | 5 |
| 27. | 'I feel anxious about receiving my diabetic eye screening results' |  |  |  |  |
|  | 1 | 2 | 3 | 4 | 5 |
| 28. | 'Diabetic eye screening appointments take up too much time in my day' |  |  |  |  |
|  | 1 | 2 | 3 | 4 | 5 |
| 29. | 'Diabetic eye screening appointment letters are sent too far in advance' |  |  |  |  |
|  | 1 | 2 | 3 | 4 | 5 |
| 30. | 'I would like more support and information following my diabetic eye screening results' |  |  |  |  |
|  | 1 | 2 | 3 | 4 | 5 |

Please describe any other factors which influence your attendance at diabetic eye screening which we have not covered

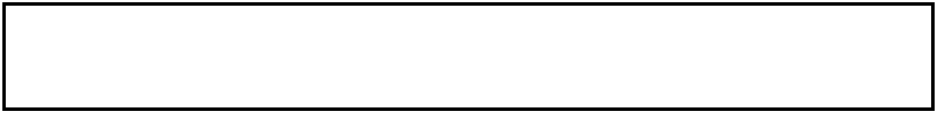

Please provide a contact e-mail address so we can include you in the prize draw

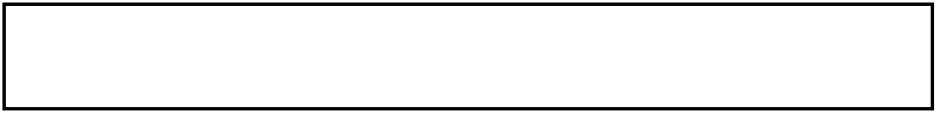

**Thank you for taking the time to complete this survey**

### S3: Rapid Digital Survey. Free text comments regarding other factors influencing DRS attendance

Q23

Please describe any other factors which influence your attendance at diabetic eye screening which we have not covered

#### COVID

- Epidemic virus
- Covid pandemic
- Last appointment later due to Covid

#### IMPACT OF EYEDROPS/TRANSPORT ISSUES

- Not able to drive somewhat dependant on others
- Not being able to drive and therefore having to find transport
- They can be difficult to get to, especially as you cannot drive-in one occasion I had to get 3 buses which took nearly 2 hours-the return trip with dilated pupils wasn’t fun. They can also be inflexible-the nearest one to me only does Tuesday mornings-ok so now I have to take a full days annual leave (I can’t read with dilated pupils so no use me going in for the afternoon) and as I need a lift to get there my husband also has to use annual leave. Also no one will tell you anything-what can you see? Is it worse than last year? Is it better? They just say wait for the results and 6 weeks of anxiety later you get a generic letter and leaflet with a grade on it but no details-fine it’s background but is it better/worse what is going on with my eyes?! Oh and my diabetologist can’t view the results, he relays on me to tell them the grading. Screening feels like it’s not for my benefit or my care but to meet a target.
- It would be easier if it was easier to get to. With the drops you can’t drive but it’s also hard to then see where the train is. The drops knock me out for the rest of the day and really affect work and everything
- Locations easy to access on public transport
- Just that your eyes stay bluey for the whole day so it almost means you have to have the rest of the day off work if you have to read things as part of your job like I do. Would be better if they did eye screens in the evening so the drops water off when you’re not trying to work.
- I much prefer having the screening during the evening during winter as the sunlight really hurts my eyes
- Travel distance and routes
- Sometimes it’s hard someone to attend the appointment with me. Which can make attendance more difficult
- Distance of screening location
- Un knowledgeable screening staff saying everything looks fine then getting a letter stating background retinopathy is very disheartening. Hard to get time off work for appointments. Horrible always being surrounded by old people in the waiting room - would LOVE a clinic just for younger people, helps to meet people in situations like mine. I have a huge fear of anything going in my eye so am terrified of any potential treatment needed. Some staff being too rigid on guidelines around the eye drops, my pupils are always huge and some staff don’t give me the drops and some say they have to even though I don’t need them just because I’m over 25

#### APPOINTMENT FEXIBILITY

- I haven’t received any appointments
- I am supposed to have my eyes screened annually but have only had 3 appointments in 6 years and very often my mum has to phone and ask for an appointment. I have little confidence in the screening program.
- Un knowledgeable screening staff saying everything looks fine then getting a letter stating background retinopathy is very disheartening. Hard to get time off work for appointments. Horrible always being surrounded by old people in the waiting room - would LOVE a clinic just for younger people, helps to meet people in situations like mine. I have a huge fear of anything going in my eye so am terrified of any potential treatment needed. Some staff being too rigid on guidelines around the eye drops, my pupils are always huge and some staff don’t give me the drops and some say they have to even though I don’t need them just because I’m over 25
- Try to give slots for weekends
- I’m really unorganised and struggle to book time off work and then end up forgetting the appointment entirely. Possibly related to dyslexia.
- Un knowledgeable screening staff saying everything looks fine then getting a letter stating background retinopathy is very disheartening. Hard to get time off work for appointments. Horrible always being surrounded by old people in the waiting room - would LOVE a clinic just for younger people, helps to meet people in situations like mine. I have a huge fear of anything going in my eye so am terrified of any potential treatment needed. Some staff being too rigid on guidelines around the eye drops, my pupils are always huge and some staff don’t give me the drops and some say they have to even though I don’t need them just because I’m over 25
- They can be difficult to get to, especially as you cannot drive-in one occasion I had to get 3 buses which took nearly 2 hours-the return trip with dilated pupils wasn’t fun. They can also be inflexible-the nearest one to me only does Tuesday mornings-ok so now I have to take a full days annual leave (I can’t read with dilated pupils so no use me going in for the afternoon) and as I need a lift to get there my husband also has to use annual leave. Also no one will tell you anything-what can you see? Is it worse than last year? Is it better? They just say wait for the results and 6 weeks of anxiety later you get a generic letter and leaflet with a grade on it but no details-fine it’s background but is it better/worse what is going on with my eyes?! Oh and my diabetologist can’t view the results, he relays on me to tell them the grading. Screening feels like it’s not for my benefit or my care but to meet a target.

#### ISSUES WITH RESULTS

- Last time my results weren’t sent to me and I had to chase for them. Results need to be provided in a timely fashion.
- I always receive letters that are extremely distressing and usually on a weekend when I cannot call anyone. Such as, “if things continue you will go blind in one eye” it is so scary and that is what I dread about these appointments.
- I was notified that I had some eye damage-but it didn’t need to be treated at this point-no one contacted me or explained the letter.
- Lack of guidance on how important it is, how to look after the eyes, nervousness of results
- They can be difficult to get to, especially as you cannot drive-in one occasion I had to get 3 buses which took nearly 2 hours-the return trip with dilated pupils wasn’t fun. They can also be inflexible-the nearest one to me only does Tuesday mornings-ok so now I have to take a full days annual leave (I can’t read with dilated pupils so no use me going in for the afternoon) and as I need a lift to get there my husband also has to use annual leave. Also no one will tell you anything-what can you see? Is it worse than last year? Is it better? They just say wait for the results and 6 weeks of anxiety later you get a generic letter and leaflet with a grade on it but no details-fine it’s background but is it better/worse what is going on with my eyes?! Oh and my diabetologist can’t view the results, he relays on me to tell them the grading. Screening feels like it’s not for my benefit or my care but to meet a target.

#### INTERACTIONS WITH SCREENING STAFF

- To be honest I find it kind of pointless as my Opticians maintains a better, easier access for support and help if I feel there is a problem, also they explain the results the doctors at the screen don’t tell me anything.
- Un knowledgeable screening staff saying everything looks fine then getting a letter stating background retinopathy is very disheartening. Hard to get time off work for appointments. Horrible always being surrounded by old people in the waiting room - would LOVE a clinic just for younger people, helps to meet people in situations like mine. I have a huge fear of anything going in my eye so am terrified of any potential treatment needed. Some staff being too rigid on guidelines around the eye drops, my pupils are always huge and some staff don’t give me the drops and some say they have to even though I don’t need them just because I’m over 25
- They can be difficult to get to, especially as you cannot drive-in one occasion I had to get 3 buses which took nearly 2 hours-the return trip with dilated pupils wasn’t fun. They can also be inflexible-the nearest one to me only does Tuesday mornings-ok so now I have to take a full days annual leave (I can’t read with dilated pupils so no use me going in for the afternoon) and as I need a lift to get there my husband also has to use annual leave. Also no one will tell you anything-what can you see? Is it worse than last year? Is it better? They just say wait for the results and 6 weeks of anxiety later you get a generic letter and leaflet with a grade on it but no details-fine it’s background but is it better/worse what is going on with my eyes?! Oh and my diabetologist can’t view the results, he relays on me to tell them the grading. Screening feels like it’s not for my benefit or my care but to meet a target.

#### FEAR OF LOSING VISION

- My mother In law lost her eye due to diabetic
- Fear of sight problems and loss
- Nothing else just that its important to keep on top of check-ups to avoid complications in the future with my eyes
- Know someone who suffered diabetic retinopathy!

#### OTHER

- In pregnancy I have to attend an eye hospital retinal screening appointment every 8 weeks until 2 months after delivery.
- So far the diabetic screening where I attended is very efficient

